# Implications of the COVID-19 pandemic on eliminating trachoma as a public health problem

**DOI:** 10.1101/2020.10.26.20219691

**Authors:** Seth Blumberg, Anna Borlase, Joaquin M Prada, Anthony W Solomon, Paul Emerson, Pamela J Hooper, Michael S Deiner, Benjamin Amoah, Deirdre Hollingsworth, Travis C Porco, Thomas M Lietman

**Author notes:** Corresponding author: Thomas M Lietman, 513 Parnassus Ave., San Francisco, CA 94143.

## Abstract

**Background:** Progress towards elimination of trachoma as a public health problem has been substantial, but the COVID-19 pandemic has disrupted community-based control efforts.

**Methods:** We use a susceptible-infected model to estimate the impact of delayed distribution of azithromycin treatment on the prevalence of active trachoma.

**Results:** We identify three distinct scenarios for geographic districts depending on whether the basic reproduction number and the treatment-associated reproduction number are above or below a value of one. We find that when the basic reproduction number is below one, no significant delays in disease control will be caused. However, when the basic reproduction number is above one, significant delays can occur. In most districts a year of COVID-related delay can be mitigated by a single extra round of mass drug administration. However, supercritical districts require a new paradigm of infection control because the current strategies will not eliminate disease.

**Conclusion:** If the pandemic can motivate judicious, community-specific implementation of control strategies, global elimination of trachoma as a public health problem could be accelerated.

## Introduction

Trachoma remains a major cause of preventable blindness, particularly in sub-Saharan Africa. Substantial reduction in the global prevalence of trachoma has been achieved, but the COVID-19 pandemic has caused unprecedented disruption of public health programs that combine surveillance of disease transmission with treatment for endemic districts. Since transmission cannot be measured directly, the World Health Organization (WHO) recommends monitoring trachoma by assessing the prevalence of trachomatous inflammation—follicular (TF) in the upper tarsal conjunctiva of children aged 1-9 years.^1,2^ For elimination of trachoma as a public health problem (termed trachoma ‘control’ herein), WHO requirements include that the prevalence of TF in children be reduced to less than five percent in each formerly-endemic district. One cornerstone of trachoma control is annual mass drug administration (MDA) of azithromycin to endemic districts.^3^ The pandemic is delaying regular MDA and thus allowing possible resurgence of active trachoma in some districts.

Based on their intensity of transmission, we can categorize districts as subcritical, MDA-subcritical, or supercritical. We delineate these levels of transmission by the basic reproduction number, *R*_*0*_, and the treatment-associated reproduction number, *R*_*T*_. We define *R*_*0*_ as the mean number of secondary infections each infection causes in the absence of MDA or immunity of close contacts.^4^ We define *R*_*T*_ to be the mean number of new infections caused by each case in a district that is receiving annual, community-wide MDA with azithromycin. We define subcritical districts to have *R*_*0*_ < 1 and thus control of trachoma would be anticipated regardless of MDA. We define MDA-subcritical districts as those with *R*_*0*_ > 1, but *R*_*T*_ < 1. These districts are progressing towards control, but only because of ongoing annual MDA. Finally, we define supercritical districts as those where both *R*_*0*_ and *R*_*T*_ are greater than one. These districts are not expected to achieve control, despite ongoing annual MDA. Our definitions of subcritical, MDA-subcritical and supercritical transmission roughly correspond to how other manuscripts describe hypoendemic, mesoendemic, and hyperendemic settings respectively.^5^ Here we utilize a mathematical model of trachoma transmission to evaluate how the disruption of MDA due to the COVID-19 pandemic may delay progress towards trachoma control.

## Methods

To simulate the prevalence of trachoma infection and estimate the delay in control caused by the COVID-19 pandemic we developed a simple model for trachoma transmission among children. We assume that children form a core group for transmission, and that transmission from adults to children is negligible.^6^ Children can either be susceptible to or actively infected with the strains of Chlamydia trachomatis that cause trachoma.^7,8^ Consistent with the classical susceptible-infected model and similar to prior studies, infection occurs at a rate that is proportional to the product of the fraction of children that are susceptible to infection and the fraction of children infected.^9,10^ Recovery from infection occurs at a rate that is proportional to the prevalence of infection. Once an infection clears, children become susceptible to infection again. Whenever a district receives a round of MDA, the number of infected people is reduced by the ‘overall MDA efficacy’, which we define as the product of azithromycin coverage and the probability that azithromycin clears infection from an individual. For Figures 1-3, we assume that the average duration of infection for children is six months, the antibiotic coverage is 80% and the azithromycin clearance is 87.5%^10^

**Figure 1:**
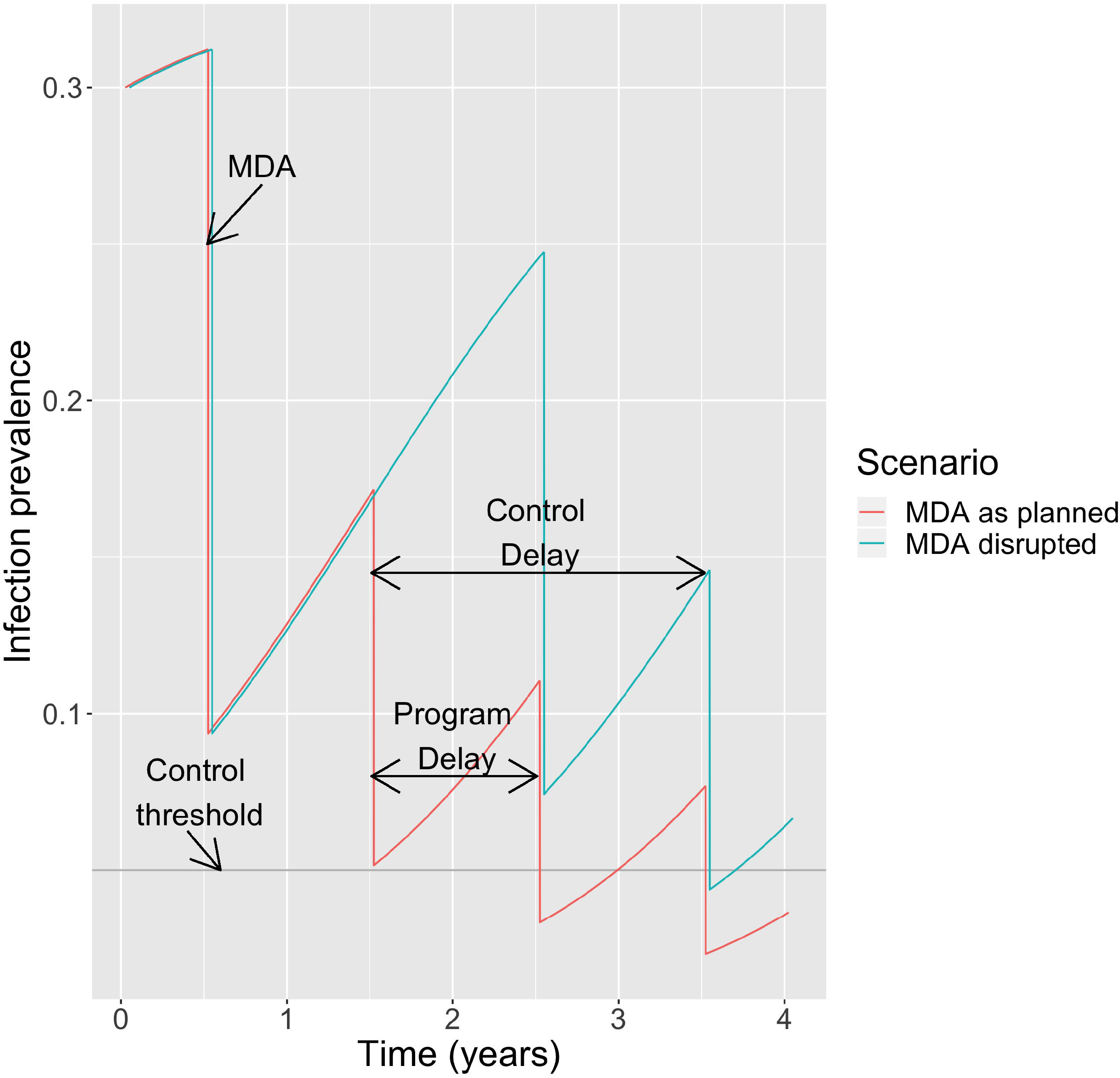
Schematic of model. The ‘MDA as planned’ scenario involves periods of exponential growth of prevalence, punctuated by uniformly spaced reduction due to annual MDA. The ‘MDA disrupted’ scenario includes one missed round of MDA at year 1.5. Infection prevalence represents the proportion of children age 1-9 with current infection. Note that infection prevalence is distinct from the clinical manifestation of trachomatous inflammation—follicular. An R^0^ of 1.5 is assumed. The ‘program delay’ is the length of MDA disruption. The ‘control delay’ is the expected delay of trachoma control due to disruption of MDA. The horizontal grey line represents the 0.05 benchmark used by WHO as part of their criteria for elimination of trachoma as a public health problem.

Based on the preceding model, trachoma control efforts are represented by periods of exponential growth (or decay) of infection that are separated by periodic reductions in infection due to annual MDA (Figure 1, red curve). The rate of exponential growth or decay is dependent on *R*_*0*_, with values of *R*_*0*_ < 1 indicating decay and *R*_*0*_ > 1 indicating growth. We define the ‘program delay’ as the time that an MDA cycle is delayed due to circumstances such as the COVID-19 pandemic (Figure 1, aqua curve). We define the ‘control delay’ as the time gap between the beginning of MDA disruption and a return to the level of infection prior to the disruption. By reflecting the additional time it will take for district-level control to be achieved, the control delay represents the overall consequence of a program delay.

We consider three scenarios for MDA. In the scenario labeled ‘MDA as planned’, one distribution of MDA occurs each year. In the scenario labeled ‘MDA disrupted’, one of the annual MDA cycles is missed. In the scenario labeled ‘MDA catch-up’, a cycle of annual MDA is missed and then a subsequent year has an additional MDA. We term this additional MDA as a ‘catch-up MDA’.

When using our model to estimate the control delay, we make the approximation that the change in prevalence between MDA time points follows an exponential curve. This is equivalent to assuming that the number of susceptible individuals is constant rather than that the entire population is constant. This assumption holds best for subcritical and MDA-subcritical districts whose prevalence was decreasing prior to MDA disruption. See supplementary text (Text S1) for more methodological details.

## Results

The impact of a one-year program delay differs depending on whether transmission is subcritical, MDA-subcritical, or supercritical (Figure 2, Table 1). In subcritical districts, the control delay is less than the program delay since prevalence is decreasing even in the absence of annual MDA. In MDA-subcritical districts, the control delay is longer than the program delay, but resumption of annual MDA cycles will put a district back on track for eventual control. An additional catch-up MDA, would return a district to its original time course, resulting in a control delay close to zero. In supercritical districts, a program delay followed by resumption of the regular MDA schedule would be roughly equivalent, but not lead to control. In contrast, a program delay followed by a catch-up MDA could accelerate control in supercritical districts.

**Table 1:** Characteristics of subcritical, MDA-subcritical, and supercritical transmission.

| | $R_0$ | $R_T$ | *Control delay* | *Catchup MDA restores progress* |
| --- | --- | --- | --- | --- |
| *Subcritical* | < 1 | < 1 | Less than program delay | Yes |
| *MDA-subcritical* | > 1 | < 1 | Greater than program delay | Yes |
| *Supercritical* | > 1 | > 1 | Not applicable as control will not be achieved | Yes |

**Figure 2:**
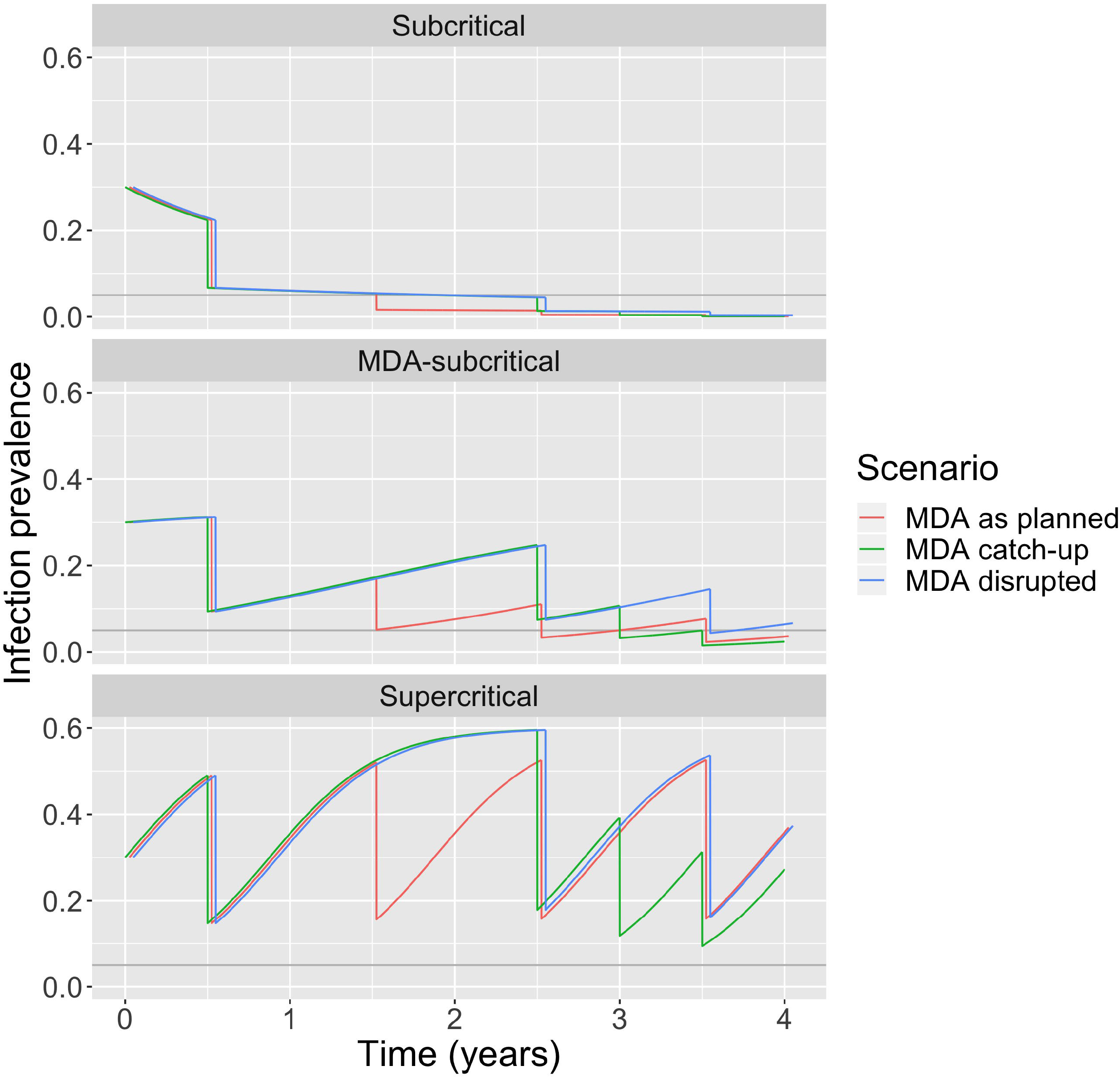
Modeling scenarios. Each panel corresponds to a different level of transmission, as defined by whether R_0_ and RT are greater or less than a value of one (Table 1). The layout of each panel is similar to Figure 1. Within each panel the ‘MDA as planned’ scenario corresponds to no disruption of annual MDA. The ‘MDA disrupted’ scenario corresponds to skipping one annual MDA cycle at year 1.5. The ‘MDA catch-up’ scenario involves giving an extra MDA at year 3, after skipping an annual MDA at year 1.5. An R_0_ of 0.95, 1.3, and 1.65 are assumed for subcritical, MDA-subcritical, and supercritical transmission respectively. For visual clarity, the time series corresponding to the scenarios are offset horizontally slightly.

Our model predicts that a program can get back on track if multiple MDA cycles are missed, provided that extra MDA cycles can be provided in subsequent years. For example, a program could get back on track after missing two annual MDA cycles if it had the resources to provide semiannual MDAs for two years before resuming annual MDA. The exact timing of catch-up MDA does not matter. For example the extra treatments could be one, three, or even six months after the scheduled annual MDA (Text S1).

Our model predicts the control delay increases as the program delay or *R*_*0*_ increases (Figure 3). For our estimates of MDA coverage and efficacy, the threshold value for *R*_*0*_ that differentiates MDA-subcritical and supercritical transmission is 1.6. As *R*_*0*_ approaches that critical point, the duration of the control delay becomes increasingly large. The theoretical delay becomes infinite for supercritical transmission. This is a reflection that supercritical districts will require new treatment strategies in order to achieve trachoma control, even had there not been a disruption in the annual MDA schedule.

**Figure 3:**
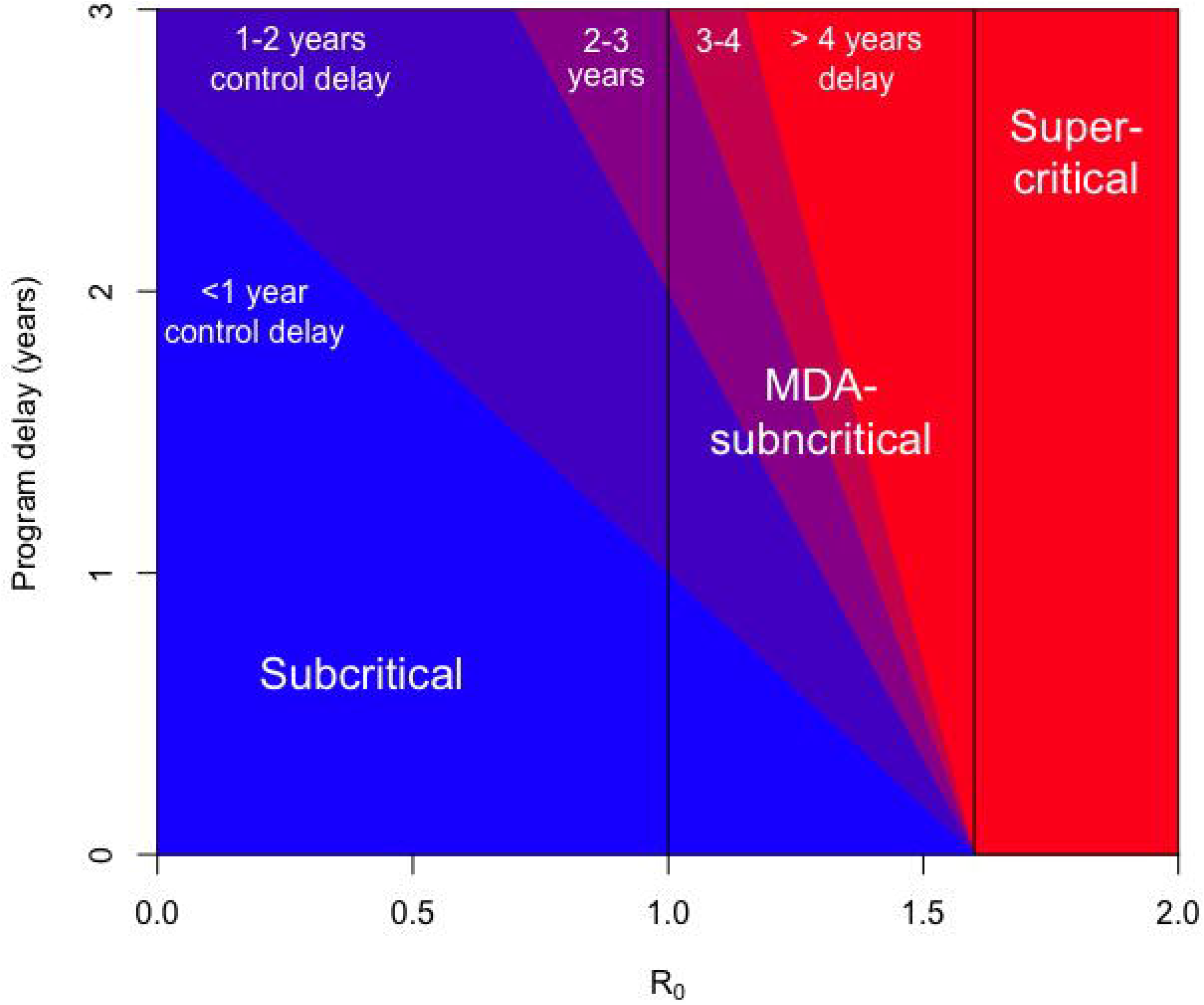
The control delay for trachoma is depicted by color, based on the R^0^ of the district and the program delay for the administration of MDA. The underlying model assumes annual MDA leads to a 70% decrease of trachoma incidence in the immediate post-treatment interval. The terms subcritical (R^0^ < 1), MDA-subcritical (1 < R^0^ < 1.6), and supercritical (R^0^ > 1.6) refer to conditions in which infection is expected to be self-limited, requires annual MDA in order for control targets to be achieved, or requires a new paradigm of treatment for eventual control, respectively. The different categories of transmission are demarcated by vertical black lines. Although our focus is on how the COVID-19 pandemic impacts trachoma control, the underlying model can be applied to a variety of diseases and program delay scenarios.

Although the prevalence of TF in any given year does not provide a direct measurement of *R*_*0*_ or *R*_*T*_, the trend of TF over time is related to these values.^11–14^ An important implication for TF surveillance is that our model of infection prevalence does not reflect the time course of TF itself. This is because TF is not a perfect marker of current infection, but rather a lagging indicator caused by the inflammatory response to infection.^15,16^ Thus, TF prevalence surveys may not register the impact of decreased infection prevalence for 6-12 months.^13,14,17,18^

Estimates of the duration of infection in children have ranged from less than ten weeks to over a year.^9,10^ In addition the overall efficacy of MDA to decrease infection in the immediate post-treatment period is unknown. Sensitivity analyses show that for subcritical districts, the control delay increases as the duration of infection increases (Figure S1, top panel). In contrast, for MDA-subcritical districts, the control delay decreases as the duration of infection increases. The same qualitative features can be seen for the impact of the overall MDA efficacy on the control delay (Figure S1, bottom panel).

## Discussion

Our model provides a quantitative perspective on how trachoma control is impacted by MDA disruption as a consequence of the COVID-19 pandemic. Interpretation of our results in the context of findings from randomized control trials (RCTs), provides a framework for strategizing the effective distribution of future MDA. Recommended strategies for maintaining progress towards trachoma control vary based on whether a region is subcritical, MDA-subcritical, or supercritical.

In subcritical districts, infection is difficult to detect, and RCTs have shown that trachoma may be controllable without multiple rounds of MDA.^19–21^ The empirical observation of self-contained transmission in subcritical districts is consistent with our model’s prediction that a program delay in MDA will not lead to a noticeable delay for district-level control. The rare apparent resurgence reported from a few subcritical districts may be attributable to a combination of misclassification and measurement error.^8,22,23^

In MDA-subcritical districts, our model suggests that the delay to achieving control will be longer than the program delay. Our model is also consistent with other modeling studies that indicate a missed cycle of MDA can be mitigated by a single catch-up MDA.^5,24^ In addition, the exact timing of the catch-up MDA does not significantly affect the performance.^25^

In supercritical districts, the control delay can vastly exceed any delay due to COVID-19. This finding aligns with prior trials and models that show infection returns more rapidly after MDA in supercritical districts.^8,26,27^ Some of these districts may eventually head towards control due to subtle reductions in disease transmission caused by changes in socioeconomic factors. However, in the limited number of districts where TF remains over 30% after a decade of MDA, our model suggests that transmission is so efficient that annual MDA will not lead to control even if there were no disruption to azithromycin distribution. Thus, alternative strategies need to be considered for control to be achieved in the near future.^28^

To address the need for adjunctive treatment strategies, much emphasis has been placed on more frequent administration of MDA. The success of this approach has been demonstrated with a RCT that compared quarterly MDA for children to annual MDA.^29^ This trial also supported the result of models that suggest children are the core group for transmission, so that adjunctive treatment for adults is unnecessary.^8^ Meanwhile, the predictions of other models of more frequent MDA have not shown good agreement with trial data. For instance, one model predicted that a second MDA given soon after the first would be especially effective at clearing infection^30^. However, this finding has yet to be verified empirically. In fact, three rounds of MDA with high antibiotic coverage in three weeks did not prevent infection from returning in the Egyptian arm of the ‘Azithromycin in control of trachoma’ study.^31^ Additional models predict that repeated biannual distribution will reduce infection more rapidly than annual distribution, but trials have not provided compelling evidence to support this prediction.^32–34^ Another adjunctive strategy for trachoma control involves improving water, sanitation, and hygiene (WASH). Models predict that if WASH measures were able to reduce transmission by 50%, there would be clear benefit.^30^ However, no trial has yet proven that WASH offers any benefit over annual MDA alone. Of note, some of the studies that failed to show a significant impact of adjunctive treatment may have been underpowered since they were conducted prior to the wide availability of sensitive nucleic acid amplification tests for identifying current infection.

Limitations of our estimate of control delay include approximations that may bias the estimate of the control delay. First, our analysis ignores how the number of susceptible individuals saturates as the prevalence of infection increases (Figure 2). Although this may be reasonable as we approach control, this approximation overestimates the between-MDA growth of the infected population in high-prevalence settings. That is, the true control delay may actually be shorter than our model’s prediction (as seen by how the ‘MDA catch-up’ is slightly better than ‘MDA as planned’ in the MDA-subcritical panel of Figure 2). In addition, the underlying assumptions of our transmission model ignore many important aspects of trachoma pathophysiology and epidemiology. These include the heterogeneity in transmission due to variable bacterial load, heterogeneity in susceptibility due to variable host immunity, and heterogeneity in contact among the population. These factors might lead to a control delay longer or shorter than the model prediction. A final consideration is that our model assumes instantaneous delivery of MDA, but logistical programmatic barriers can cause delays in drug delivery within districts. Some of these limitations can be addressed by stochastic models that incorporate more than one state of infection.^5,24^

Despite the limitations of the model, we hope our results can be useful for stakeholders involved in trachoma control (Table 2).^35^ From a policy perspective, our results provide reassurance that the resumption of trachoma control is feasible. Further work is needed to classify districts as MDA-subcritical versus supercritica; so that district-level distribution of MDA can be optimized. Catch-up treatments may be beneficial in MDA-subcritical districts, but the exact timing will likely not matter. In addition, maintenance of pre-pandemic MDA coverage and efficacy should be prioritized over early resumption of MDA with suboptimal coverage. Meanwhile, supercritical districts will require an alternate treatment strategy to annual MDA.

**Table 2:** Summary of Policy-Relevant Items for Reporting Models in Epidemiology of Neglected Tropical Diseases.^35^. BMGF = Bill and Melinda Gates Foundation. ITI = International Trachoma Initiative

| Policy Relevant principle | Application to manuscript | Location of specific detail |
| --- | --- | --- |
| Stakeholder engagement | Study inspired by NTD consortium which maintains direct communication with WHO, BMGF, ITI and other stakeholders | Discussion |
| Complete model documentation | A complete description of our model is provided. Code is available in a GitHub repository | Methods / supplement<br>https://github.com/sblumberg/trachoma---COVID_impact |
| Complete description of data used | Model is parameterized by previously published results focused on the transmission dynamics of trachoma. | Methods |
| Communicating uncertainty | Assumptions of the model that lead to uncertainty in the results are described<br>Sensitivity analyses for the duration of infectious period and overall MDA efficacy are presented | Discussion<br>Supplemental figure |
| Testable model outcomes | An alternative model is presented in this special collection. Given the unprecedented nature of the COVID-19 pandemic, no data are currently available for model validation. | See Borlase et al.<sup>5</sup> |

## Conclusion

When considering the impact of the COVID-19 pandemic on efforts to control trachoma, districts can be classified as subcritical, MDA-subcritical, or supercritical. In subcritical districts, no significant delay in achieving control goals is anticipated. In MDA-subcritical districts, control after a one-year program delay can be achieved by either extending the duration of annual MDA distribution or by providing an additional catch-up round of MDA. Meanwhile, supercritical districts will require adjunctive treatments in order to reach control milestones. Although models have endorsed a variety of adjunctive treatments in supercritical districts, quarterly MDA for children remains the only intervention shown to be statistically superior in field trials.^29^ The COVID-19 pandemic may offer the opportunity to reassess strategies to achieve trachoma control in these districts.

## Data Availability

This is a modeling paper that does not present new data. All methods and parameter values are described within the paper. All code is available at:
https://github.com/sblumberg/trachoma---COVID_impact

https://github.com/sblumberg/trachoma---COVID_impact

## Authors’ contributions

This manuscript arose from a virtual meeting of the NTD Modeling Consortium for which all authors contributed analyses and discussions. SB, TML, and TCP wrote the first draft. All authors edited the manuscript and approved the final draft.6 The authors alone are responsible for the views expressed in this article and they do not necessarily represent the views, decisions or policies of the institutions with which they are affiliated.

## Acknowledgments

We thank Simon J Brooker for helping to motivate this research and for his valuable feedback on preliminary work.

## Supplement

Figure S1: Sensitivity analysis for how the control delay is affected by the duration of the infectious period (top) and the overall MDA efficacy (bottom). For both panels, the program delay is assumed to be one year. In the top panel, the overall MDA efficacy is assumed to 70%. In the bottom panel, the average duration of the infection period is assumed to be 26 weeks. The vertical black line differentiates subcritical transmission from MDA-subcritical transmission.

Text S1: Technical description of how control delay is calculated

## References

1. Meeting for the development of guidelines for assessment of the elimination of blinding trachoma. Presented at the: November 8, 2001; Geneva, Switzerland: World Health Organization, Prevention of Blindness & Deafness.

2. Thylefors B, Dawson CR, Jones BR, West SK, Taylor HR. A simple system for the assessment of trachoma and its complications. Bull World Health Organ. 1987;65(4):477–483.

3. WHO Alliance for the Global Elimination of Trachoma. Meeting (3rd: 1998: Ouarzazate M, Deafness WP for the P of B and. Report of the third meeting of the WHO Alliance for the Global Elimination of Trachoma, Ouarzazate, Morocco, 19-20 October 1998. Published online 1999. Accessed June 29, 2020. https://apps.who.int/iris/handle/10665/65933

4. Anderson RM, May RM. Infectious Diseases of Humans: Dynamics and Control. Oxford University Press; 1991.

5. Borlase, Anna. Companion article for TRSTMH special edition,”“Invited Papers for COVID 19 NTD Programme Interruption” - In prep.

6. Solomon AW, Holland MJ, Burton MJ, et al. Strategies for control of trachoma: observational study with quantitative PCR. Lancet Lond Engl. 2003;362(9379):198–204. doi:10.1016/S0140-6736(03)13909-8

7. Lietman TM, Deiner MS, Oldenburg CE, Nash SD, Keenan JD, Porco TC. Identifying a sufficient core group for trachoma transmission. PLoS Negl Trop Dis. 2018;12(10):e0006478. doi:10.1371/journal.pntd.0006478

8. Lietman T, Porco T, Dawson C, Blower S. Global elimination of trachoma: how frequently should we administer mass chemotherapy?Nat Med. 1999;5(5):572–576. doi:10.1038/8451

9. Grassly NC, Ward ME, Ferris S, Mabey DC, Bailey RL. The natural history of trachoma infection and disease in a Gambian cohort with frequent follow-up. PLoS Negl Trop Dis. 2008;2(12):e341. doi:10.1371/journal.pntd.0000341

10. Ray KJ, Lietman TM, Porco TC, et al. When can antibiotic treatments for trachoma be discontinued? Graduating communities in three African countries. PLoS Negl Trop Dis. 2009;3(6):e458. doi:10.1371/journal.pntd.0000458

11. Bird M, Dawson CR, Schachter JS, et al. Does the diagnosis of trachoma adequately identify ocular chlamydial infection in trachoma-endemic areas?J Infect Dis. 2003;187(10):1669–1673. doi:10.1086/374743

12. Keenan JD, Lakew T, Alemayehu W, et al. Clinical activity and polymerase chain reaction evidence of chlamydial infection after repeated mass antibiotic treatments for trachoma. Am J Trop Med Hyg. 2010;82(3):482–487. doi:10.4269/ajtmh.2010.09-0315

13. Koukounari A, Moustaki I, Grassly NC, et al. Using a Nonparametric Multilevel Latent Markov Model to Evaluate Diagnostics for Trachoma. Am J Epidemiol. 2013;177(9):913–922. doi:10.1093/aje/kws345

14. Liu F, Porco TC, Amza A, et al. Short-term forecasting of the prevalence of clinical trachoma: utility of including delayed recovery and tests for infection. Parasit Vectors. 2015;8:535. doi:10.1186/s13071-015-1115-8

15. Solomon AW, Foster A, Mabey DCW. Clinical examination versus Chlamydia trachomatis assays to guide antibiotic use in trachoma control programmes. Lancet Infect Dis. 2006;6(1):5–6; author reply 7-8. doi:10.1016/S1473-3099(05)70304-2

16. Wright HR, Taylor HR. Clinical examination and laboratory tests for estimation of trachoma prevalence in a remote setting: what are they really telling us?Lancet Infect Dis. 2005;5(5):313–320. doi:10.1016/S1473-3099(05)70116-X

17. Bird M, Dawson C, Miao Y, Schachter J, Lietman T. Does the Clinical Exam Adequately Identify Ocular Chlamydial Infection in Trachoma?Invest Ophthalmol Vis Sci. 2002;43(13):3061–3061.

18. Keenan JD, Lakew T, Alemayehu W, et al. Slow resolution of clinically active trachoma following successful mass antibiotic treatments. Arch Ophthalmol Chic Ill 1960. 2011;129(4):512–513. doi:10.1001/archophthalmol.2011.46

19. Harding-Esch EM, Sillah A, Edwards T, et al. Mass treatment with azithromycin for trachoma: when is one round enough? Results from the PRET Trial in the Gambia. PLoS Negl Trop Dis. 2013;7(6):e2115. doi:10.1371/journal.pntd.0002115

20. Jimenez V, Gelderblom HC, Flueckiger RM, Emerson PM, Haddad D. M ass Drug Administration for Trachoma: How Long Is Not Long Enough? PLoS Negl Trop Dis. 2015;9(3):e0003610. doi:10.1371/journal.pntd.0003610

21. Wilson N, Goodhew B, Mkocha H, et al. Evaluation of a Single Dose of Azithromycin for Trachoma in Low-Prevalence Communities. Ophthalmic Epidemiol. 2019;26(1):1–6. doi:10.1080/09286586.2017.1293693

22. Godwin W, Prada JM, Emerson P, et al. Trachoma Prevalence After Discontinuation of Mass Azithromycin Distribution. J Infect Dis. Published online February 13, 2020. doi:10.1093/infdis/jiz691

23. Lietman TM, Gebre T, Ayele B, et al. The epidemiological dynamics of infectious trachoma may facilitate elimination. Epidemics. 2011;3(2):119–124. doi:10.1016/j.epidem.2011.03.004

24. Toor, Jaspreet et al. Predicted impact of COVID-19 on neglected tropical disease programs and the opportunity for innovation. Clin Infect Dis. In press.

25. Gao D, Lietman TM, Dong C-P, Porco TC. Mass drug administration: the importance of synchrony. Math Med Biol J IMA. 2017;34(2):241–260. doi:10.1093/imammb/dqw005

26. Melese M, Chidambaram JD, Alemayehu W, et al. Feasibility of eliminating ocular Chlamydia trachomatis with repeat mass antibiotic treatments. JAMA. 2004;292(6):721–725. doi:10.1001/jama.292.6.721

27. Lakew T, House J, Hong KC, et al. Reduction and return of infectious trachoma in severely affected communities in Ethiopia. PLoS Negl Trop Dis. 2009;3(2):e376. doi:10.1371/journal.pntd.0000376

28. Lietman TM, Pinsent A, Liu F, Deiner M, Hollingsworth TD, Porco TC. Models of Trachoma Transmission and Their Policy Implications: From Control to Elimination. Clin Infect Dis Off Publ Infect Dis Soc Am. 2018;66(Suppl 4):S275–S280. doi:10.1093/cid/ciy004

29. House JI, Ayele B, Porco TC, et al. Assessment of herd protection against trachoma due to repeated mass antibiotic distributions: a cluster-randomised trial. Lancet Lond Engl. 2009;373(9669):1111–1118. doi:10.1016/S0140-6736(09)60323-8

30. Pinsent A, Burton MJ, Gambhir M. Enhanced antibiotic distribution strategies and the potential impact of facial cleanliness and environmental improvements for the sustained control of trachoma: a modelling study. BMC Med. 2016;14(1):1–10. doi:10.1186/s12916-016-0614-6

31. Schachter J, West SK, Mabey D, et al. Azithromycin in control of trachoma. Lancet Lond Engl. 1999;354(9179):630–635. doi:10.1016/S0140-6736(98)12387-5

32. Melese M, Alemayehu W, Lakew T, et al. Comparison of annual and biannual mass antibiotic administration for elimination of infectious trachoma. JAMA. 2008;299(7):778–784. doi:10.1001/jama.299.7.778

33. Gebre T, Ayele B, Zerihun M, et al. Comparison of annual versus twice-yearly mass azithromycin treatment for hyperendemic trachoma in Ethiopia: a cluster-randomised trial. Lancet Lond Engl. 2012;379(9811):143–151. doi:10.1016/S0140-6736(11)61515-8

34. Amza A, Kadri B, Nassirou B, et al. A Cluster-Randomized Trial to Assess the Efficacy of Targeting Trachoma Treatment to Children. Clin Infect Dis Off Publ Infect Dis Soc Am. 2017;64(6):743–750. doi:10.1093/cid/ciw810

35. Behrend MR, Basáñez M-G, Hamley JID, et al. Modelling for policy: The five principles of the Neglected Tropical Diseases Modelling Consortium. PLoS Negl Trop Dis. 2020;14(4):e0008033. doi:10.1371/journal.pntd.0008033

