## Supplemental text - S1 for "Implications of the COVID-19 pandemic on eliminating trachoma as a public health problem"

### Analysis

Technical details: Calculation of control delay

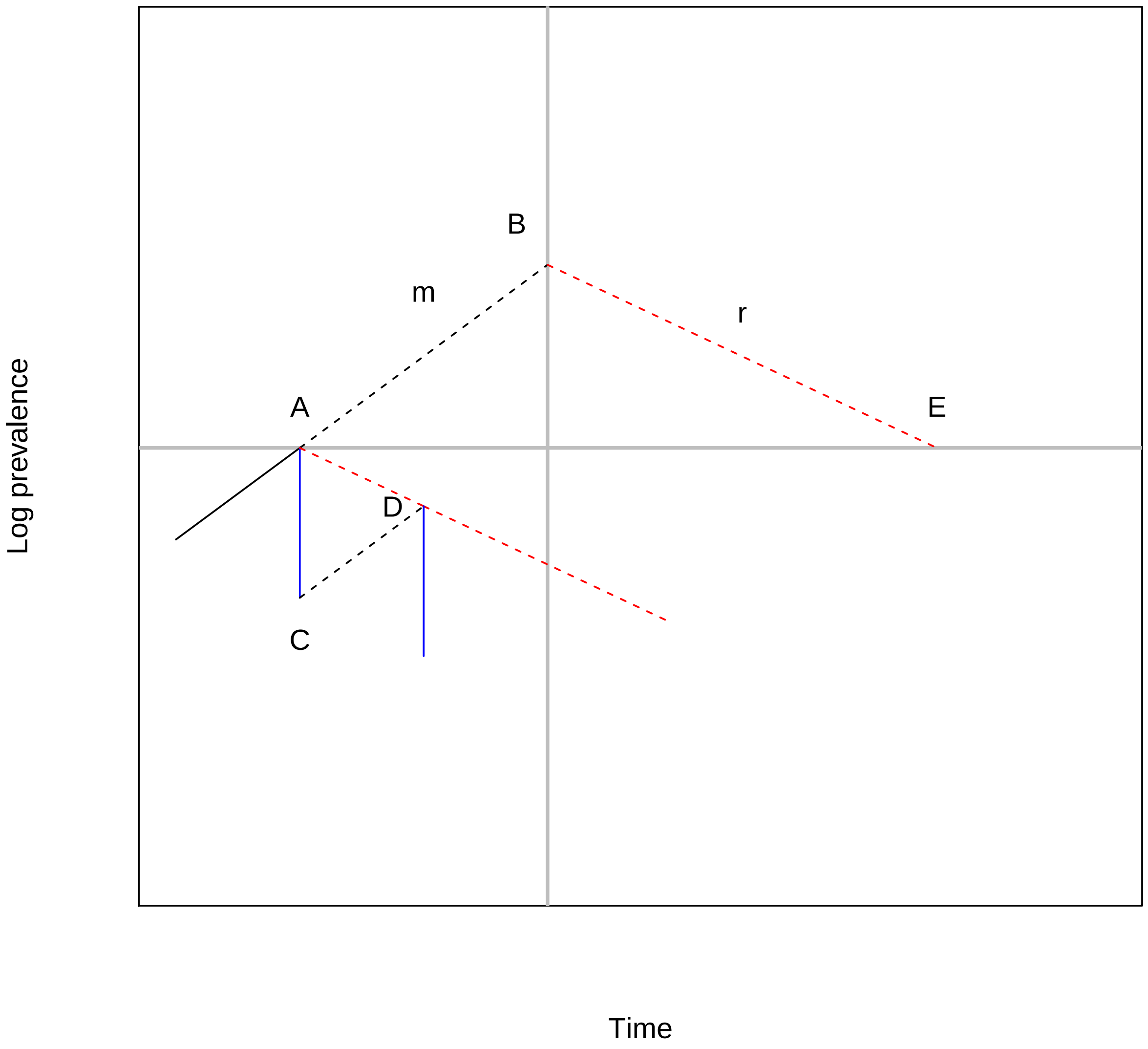


The figure illustrates relevant phenomena. Point A is the prevalence at the time of program interruption.

The blue vertical line shows what would happen if a treatment were to occur at that time—a drop in prevalence to point C, after which prevalence would resume increasing to point D, the next intended distribution. The magnitude of the drop for an intervention, i.e., the length of the blue line, is denoted $\theta$.

$$\theta = -\log(1 - ce)$$

where $c$ is coverage and $e$ is efficacy.

The red dashed line is the overall decline rate in the presence of the regular program.

In the absence of treatment, prevalence is assumed to increase until point B, with slope $m$. The time between B and A is the **program delay** due to COVID-19. At point B, the overall rate is assumed to change at average rate $r$, following the red dashed line. By point E, the prevalence has returned to the value at A. The additional time from B to E is the **catchup time**, so the total **control delay** is the program delay plus the catchup time.

Let $t_0$ denote the program delay, and $t_1$ the catchup time. The control delay is then $t_0 + t_1$. Since $mt_0 = rt_1$, the control delay $S$ is

$$S = t_0 + \frac{m}{r}t_0 = t_0\big(1 + \frac{m}{r}\big).$$

Here $r$ is the magnitude of the slope. The assumption is that the slope is strictly negative—the program is able to eventually control the disease.

Let $T$ be the intertreatment interval under normal conditions (one year). Then the difference in height of point D relative to point A must be $mT - \theta$. Thus,

$$r = \frac{\theta - mT}{T}$$

and

$$S = t_0 \frac{\theta}{\theta - mT}$$

The minus sign in the denominator is not a problem; if $r = 0$, it is never possible to return to the conditions at point A since the program is incapable of reducing prevalence under this assumption.

Finally $m = (R_0 - 1)\gamma$, so

$$S = t_0 \frac{\theta}{\theta - (R_0 - 1)\gamma T}.$$
