## Supplementary figures and images for "Implications of the COVID-19 pandemic on eliminating trachoma as a public health problem"

### Supplemental figure - S1

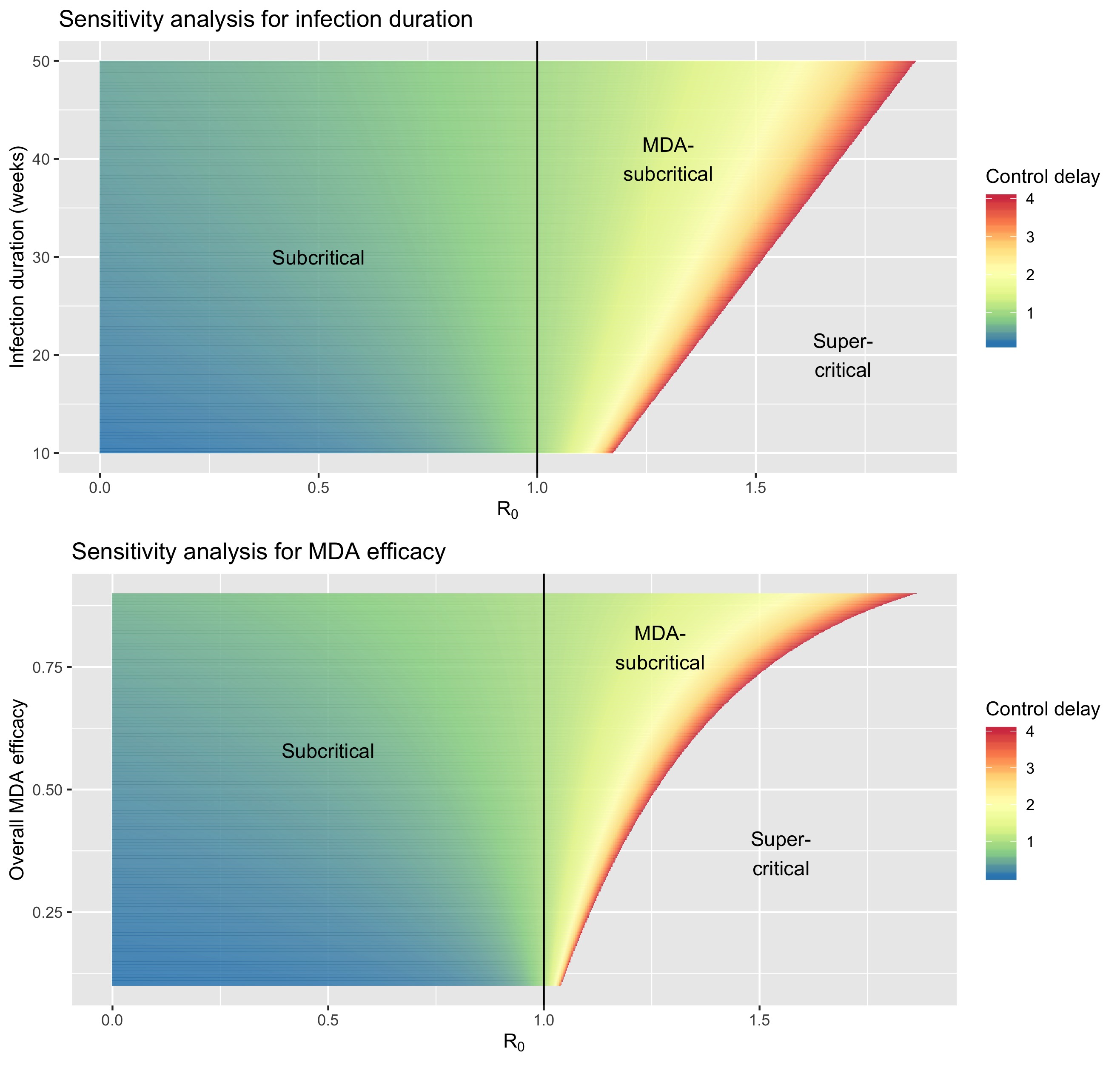
